# Economic Burden of Glaucoma Among Patients Visiting University of Gondar Comprehensive Specialized Hospital, Tertiary Eye Care and Training Centre, Northwest Ethiopia: A Cross-Sectional Study

**DOI:** 10.64898/2026.11.17.26368360

**Authors:** Simegnew Nibret, Fisseha Admassu, Tesfahun Ejigu, Banchilay Addis, Getasew Ayichew

## Abstract

Glaucoma is a chronic illness that can create a significant financial burden for patients. A hospital-based cross-sectional study was conducted to assess the economic burden of glaucoma among patients at the University of Gondar Tertiary Eye Care and Training Centre. Consecutive sampling was used, and 325 patients who had taken antiglaucoma medication for at least six months participated in the study. Data were collected using a structured questionnaire; after a proper checkup, the data were entered into EpiData version 4.7 and exported to Stata version 17 for further analysis. Descriptive statistics, bivariate screening, and final multiple linear regression after log transformation were performed to assess outcome variables as well as the relationship between the total cost and independent variables. The mean age of the participants was 61 (range: 18–90). Males accounted for 60.31%. The most common type of glaucoma was POAG, which accounted for 67.38%, followed by pseudoexfoliative glaucoma (12.92%). The mean total cost related to glaucoma management per patient in this study was 904.75 ETB (5.86 USD) monthly. This constituted 22.1% of the participants’ mean monthly income. Total cost related to glaucoma was significantly associated with residency, health insurance coverage, laterality of the disease, number of anti-glaucoma medications, and history of glaucoma surgery. The study revealed that glaucoma had a significant effect on patients’ economic, personal, and social quality of life. It is recommended to expand the community-based health insurance system to include those anti-glaucoma drugs bought from private pharmacies.

## Introduction

Glaucoma is a group of progressive optic neuropathies that result in continuous damage to the optic nerve and subsequent distinctive morphological changes at the optic nerve head and retinal nerve fiber [1]. The prevalence of glaucoma globally is 3.54%. Primary open-angle glaucoma (POAG) is the most common type, accounting for 3.05% of cases. In 2013, it was estimated that globally, 64.3 million adults aged 40–80 years lived with glaucoma [2]. Glaucoma treatment primarily aims to reduce intraocular pressure (IOP) through medical, laser, or surgical means. The first-line treatment typically involves medications, and laser and/or surgical options can be utilized when the IOP is not well controlled [3].

Patients with glaucoma may face financial challenges as well as difficulties in carrying out routine activities [4]. A study in the United States and Australia found that glaucoma imposed substantial direct and indirect costs. Direct costs include medication, surgery, diagnostic procedures, and transportation fees, while indirect costs include economic losses from work absenteeism and diminished caregivers’ capacity to perform daily duties [5]. About 58.11% of glaucoma patients in India also had financial hardship, and non-adherence to anti-glaucoma medications was common, especially for drugs that could not be accessed in public pharmacies or were too expensive for private purchases. In addition, a large portion (37.84%) of participants were retired and financially dependent on others for their medication bills. Only 4.05% of patients had medication costs reimbursed from government health schemes, leaving most to bear the full financial load [6]. Based on the study conducted in Nigeria, about half (49.76%) of the patients’ monthly income was spent on the payment of anti-glaucoma medications, diagnostic tests, transportation, and time lost from work [7]. Similarly, in Jimma, Ethiopia, the financial burden was significant among glaucoma patients; up to 29% of their reported monthly income was spent on medications. Moreover, patients from rural areas spent up to 54.5% of their monthly income on ocular hypotensive drugs, and those who had advanced glaucoma spent a higher cost than patients with early and moderate glaucoma [4].

Glaucoma requires frequent patient visits and ongoing interventions, which can lead to financial challenges. Despite this, studies assessing the economic burdens of glaucoma in Ethiopia are limited. To the best of our knowledge, no similar study has been conducted in the study area. Therefore, this study was conducted to evaluate the economic impact of glaucoma among patients attending the University of Gondar, Comprehensive Specialised Hospital, Gondar, Ethiopia.

## Methods and Materials

### Study Area

This study was conducted at the University of Gondar, Comprehensive Specialised Hospital, Gondar City, Northwest Ethiopia. The city is located in the Central Gondar administrative zone, Amhara National Regional State, 750 km from Addis Ababa, the capital city of Ethiopia, and 180 km from Bahir Dar, the capital city of the Amhara Regional State. Founded in 1982, the Department of Ophthalmology at the University of Gondar Comprehensive Specialized Hospital (GUCSH) is one of the two tertiary eye care facilities in Northwest Ethiopia that offer comprehensive care, speciality eye care services, and undergraduate as well as postgraduate training. It has a referral link from eight zones, with a total catchment area of 14 million people [8].

### Study Design and Study Period

A hospital-based cross-sectional study was conducted from March 2025 to December 2025.

### Study Participants

All patients who underwent follow-up at the glaucoma clinic during the study period were the source population.

The study population include patients aged greater than 18 years, those who had had follow-up for at least 6 months at the glaucoma clinic during the study period, those willing to give their consent to the study, and patients without concomitant ophthalmic disorders requiring medical attention and costing them, including diabetic retinopathy, AMD, RVO, and retinal detachment, were the study population.

### Sample Size and Sampling Procedures

The sample size was calculated using the following single proportion formula: (α/2) = level of significance, which was 95% = 1.96, and p = proportion. From a study conducted in Jimma, Ethiopia, where patients spent 29% of their reported income on medications only, we assumed a p-value approaching 29% (p=0.29) [4].

q = 1 -p = 0.71, e = 5% margin of error

n= ((z a⁄2)2 p(1-p))/e2

n= ((1.96)2 0.29(1-0.29))/ (0.05)2

n = 316; assuming a 5% non-response rate, the total sample size was 332.

The total sample size attained during the study period was 325, and the consecutive sampling technique was implemented, in which all patents that fulfilled the inclusion criteria were selected for the study.

### Study Variables

The dependent variable was total cost related to glaucoma management, whereas demographic variables including age, sex, marital status, educational status, occupation, and residency; disease/glaucoma-related as well as cost-related variables like type of glaucoma, laterality, stage of glaucoma, duration of follow-up, income level, health insurance status, number of antiglaucoma medications, monthly cost of medications, history of surgery, average frequency of follow-up, and number of man-hours lost during hospital visits were independent variables.

### Operational Definitions

**Income** refers to the total value of all earnings received over a specific period, including wages, salaries, profits, interest, rental income, and other forms of compensation.

**The stage or severity** of glaucoma was determined using the Canadian Ophthalmological Society’s evidence-based clinical guidelines for adult glaucoma management [9].

**Early stage**: Defined by early signs of glaucomatous optic disc changes (e.g., cup-to-disc ratio [C/D*] < 0.65) and/or a mild visual field defect outside the central 10° (e.g., mean deviation [MD] better than –6 dB on the Humphrey visual field test).

**Moderate stage**: Defined by moderate disc abnormalities (e.g., vertical C/D* between 0.7 and 0.85) and/or moderate visual field loss outside the central 10° (e.g., MD between –6 and –12 dB on the 24-2 test on the Humphrey visual field test).

**Advanced stage**: Defined by severe optic nerve changes (e.g., C/D* > 0.9) and/or a significant visual field defect within the central 10° (e.g., MD worse than –12 dB on the Humphrey visual field test).

### Cost classifications

**Direct costs** are directly tied to the disease and its treatment. This includes medical costs (such as medications, diagnostics, and procedures), non-medical costs (such as transportation, home care, assistive devices, and informal caregiving), and time spent in treatment by patients or caregivers [10–12].

**Indirect costs** reflect the economic impact of reduced productivity due to illnesses or disabilities. These include lost work time (absenteeism), reduced working hours (part-time work), salary loss, and loss of work [10–12].

**Intangible costs** or effects refer to costs resulting from pain and suffering caused by a disease. The burden of illness on affected persons and caregivers comprises, among others, loss of well-being or loss of quality of life [10, 11].

### Data Collection Procedures and Tools

A standardized questionnaire, which was prepared after reviewing important points from different literature, was used to gather data from patients, their caregivers/parents, next of kin or guardians, and the patients’ medical charts. It has both Amharic and English forms to ensure information accuracy and consistency. The questionnaire was pretested by interviewing a sample of patients. Data were collected by the principal investigator and the team. The principal investigator provided one day of training for the data collector about the purpose of the study, data collection process, consent, and other ethical issues. Informed written consent to participate in the study was obtained from all participants.

Patients’ charts were reviewed for inclusion and exclusion criteria before collecting the necessary information, and the medical record charts of the patients from whom data were collected were marked to avoid duplication. The principal investigator assessed the completeness and consistency of the collected data each day.

### Statistical Analysis

Data were entered into EpiData version 4.7 and then exported to Stata version 17 MP for further analysis. The normality of the numerical outcomes of interest (total cost) was checked with a histogram, Shapiro-Wilk test, and skewness/kurtosis test; it was skewed to the right with a p-value < 0.05. Linearity was checked with scatter plots and found to be nonlinear. A log transformation of the total cost was applied, resulting in an improvement in these parameters, and multiple linear regression was subsequently conducted to assess the association between the dependent variable, that is, total cost, and independent variables such as sociodemographic and glaucoma-related factors and cost-related variables. Bivariate screening was performed; variables with a p-value < 0.25 were selected for multivariate analysis, in which variables with a p-value < 0.05 were considered statistically significant. Descriptive statistics were used to assess the magnitude of intangible burdens.

## Ethical Considerations

Ethical clearance was obtained from the University of Gondar School of Medicine Ethical Review Committee with reference number SOM 460/2025, and permission to proceed with this study was obtained from the Department of Ophthalmology. The study participants were recruited from March 10, 2025, to December 25, 2025. Informed written consent was obtained after a brief explanation of the study to the patients, and they were assured that they had the full right to refuse the study. The patient’s name was not recorded during information gathering; confidentiality was respected, and the study was conducted based on the Helsinki Declaration.

## Result

### Socio-demographic Profiles

A total of 325 participants were included in this study; the mean age of participants was 61 (range, 18-90), and the dominant age group was 60 to 70 (33.5%). Males accounted for 196 (60.31%). Of the participants, 98 (30.15%) were farmers, 93 (28.62%) were housewives, and 60 (18.46%) were retirees. Approximately half of the participants could not read or write (169; 52.00%). The majority of our participants (278; 85.54%) were married; 176 (54.15%) of those participants were from rural areas (Table 1).

**Table 1:**
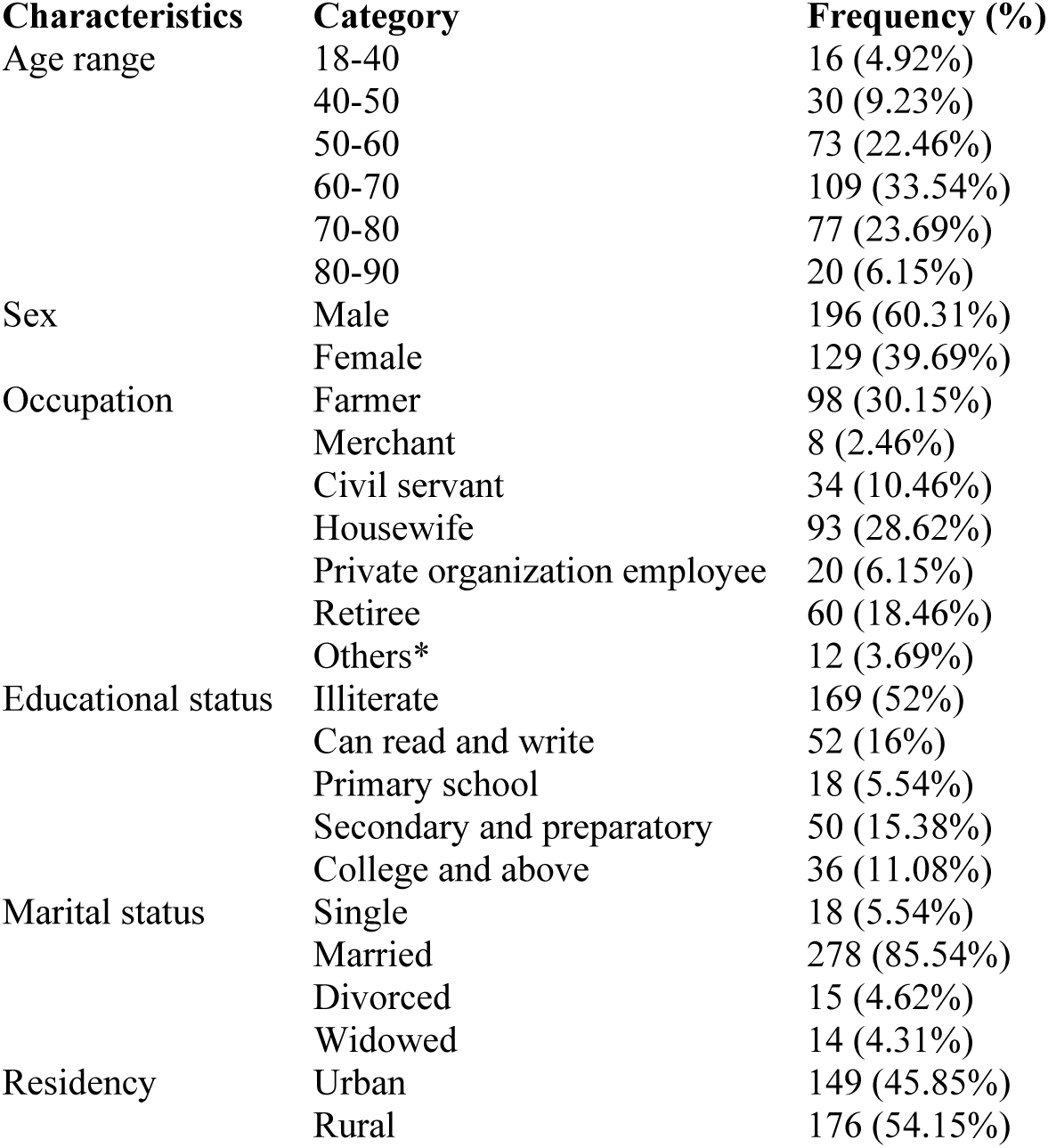
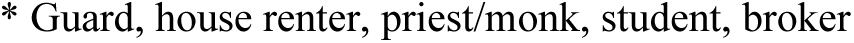
Sociodemographic Characteristics of Study Participants.

### Disease-related Characteristics

As shown in Table 2, the most common glaucoma type, which was diagnosed in two-thirds of the participants (219 (67.38%)), was POAG. Patients with advanced glaucoma had a higher proportion (184 (56.62%)). Most study participants (187 (57.54%)) were on antiglaucoma medication for both eyes. Patients who were on antiglaucoma medication for 2–5 years were predominant (128 (39.38%)). Associated ocular conditions coexisted in 63 (19.38%) glaucoma patients, and the most common was age-related immature cataract (ARIC), which was found in 43 (13.18%) of them. Associated systemic diseases were found in 92 (28.31%) participants, and the predominant diseases were HTN in 37 (11.38%) and DM in 14 (15.22%).

**Table 2:** Disease-related Characteristics.

| **Characteristics** | **Category** | **Frequency (%)** |
|---|---|---|
| Type of glaucoma | POAG | 219 (67.38%) |
|  | PXFG | 42 (12.92%) |
|  | PACG | 37 (11.38%) |
|  | NTG | 15 (4.62%) |
|  | JOAG | 10 (3.08%) |
|  | Steroid-induced | 1 (0.31%) |
|  | Secondary ACG | 1 (0.31%) |
| Stage of glaucoma | Early | 33 (10.15%) |
|  | Moderate | 108 (33.23%) |
|  | Advanced | 184 (56.62%) |
| Laterality | One eye | 138 (42.46%) |
|  | Both eyes | 187 (57.54%) |
| Duration of treatment with AGM | 6 months to 1 year | 41 (12.62%) |
|  | 1 to 2 years | 92 (28.31%) |
|  | 2 to 5 years | 128 (39.38%) |
|  | 5 or more years | 64 (19.69%) |
| Presence of other ocular diseases | Yes. | 63 (19.38%) |
|  | No | 262 (80.62%) |
|  | ARIC | 43 (13.18%) |
| If yes, which disease? | Pterygium | 2 (0.62%) |
|  | VKC | 3 (0.93%) |
|  | MGD, blepharitis | 3 (0.93%) |
|  | Others* | 12 (3.72%) |
| Associated systemic disease | Yes. | 92 (28.31%) |
|  | No | 233 (71.69%) |

Others\*: anophthalmic socket (fellow eye), corneal opacity, trachomatous trichiasis, post-OSSN excision, phthisis bulbi, dry AMD, pathologic myopia, treated optic neuritis

### Cost-related Characteristics

Approximately two-thirds of the study participants (224 (68.92%)) reported that their monthly income was less than 5000 ETB or nil. Most (59.08%) covered treatment costs primarily by themselves. The majority of study participants (269, 82.77%) had health insurance at the time of the study. Of the insured groups, medication costs were covered partly (they had to pay out-of-pocket for some of their medications) in 212 (78.81%) and fully in 56 (17.23%) subjects. Patients on monotherapy were 254 (78.15%). About 39.62% of glaucoma patients had been offered surgery, and 32.62% of them had undergone glaucoma surgical procedures such as trabeculectomy, LPI, or SLT. The usual follow-up was every 2 months in 131 (40.31%) subjects. The estimated man-hours lost during each hospital visit were predominantly 2 to 6 hours among 87 (26.77%); and 130 (40.00%) patients reported that they waited for doctors for 2 to 4 hours (see Table 3).

**Table 3:** Cost-related Characteristics.

| **Characteristics** | **Category** | **Frequency (%** |
|---|---|---|
| Monthly income in ETB | <5000 or nil | 224 (68.92%) |
|  | 5000-10000 | 62 (19.08%) |
|  | 10000-20000 | 37 (11.38%) |
|  | >=20000 | 2 (0.62%) |
| Principal payer for care services | Self | 192 (59.08%) |
|  | Family member | 85 (26.15%) |
|  | Other relatives | 1 (0.31%) |
|  | Government | 1 (0.31%) |
|  | Non-governmental organizations | 2 (0.62%) |
|  | Health insurance | 44 (13.54%) |
| Health insurance status | Yes. | 269 (82.77%) |
|  | No | 56 (17.23%) |
| Health insurance coverage for medication costs | Partly | 212 (78.81%) |
|  | Fully | 41 (15.24%) |
|  | Not at all | 16 (5.95%) |
| Number of AGMs | 1 | 254 (78.15%) |
|  | 2 | 63 (19.38%) |
|  | 3 | 8 (2.46%) |
| Offered to undergo surgery | Yes. | 129 (39.69%) |
|  | No | 196 (60.32%) |
| Undergone surgery | Yes. | 106 (32.62%) |
|  | No | 219 (67.38%) |
| Frequency of follow-up | Less than 1 month | 6 (1.85) |
|  | Every month | 72 (22.15) |
|  | Every 2 months | 131 (40.31) |
|  | Every 3 months | 116 (35.69) |
| Number of man-hours lost during each hospital visit | Less than 2 hours | 37 (11.38) |
|  | 2 to 6 hours | 87 (26.77) |
|  | 6 to 8 hours | 60 (18.46) |
|  | 8 to 16 hours | 79 (24.31) |
|  | 16 to 24 hours | 28 (8.62) |
|  | 24 to 48 hours | 31 (9.54) |
|  | More than 48 hours | 3 (0.92) |
| Man-hours lost to get a doctor | 1 hour | 11 (3.38) |
|  | 2 hours | 71 (21.85) |
|  | 2 to 4 hours | 130 (40) |
|  | more than 4 hours | 113 (34.77) |

The most commonly prescribed glaucoma drug was monotherapy with timolol (232 patients, 71.38%), followed by timolol with dorzolamide dual therapy (29 patients, 8.92%). The least costly medication in our study was timolol eye drops, with a mean monthly cost of 82.5 ETB ($0.53), whereas the costliest one was triple therapy of timolol + dorzolamide + latanoprost with a monthly cost of 2610 ETB ($16.95). Other anti-glaucoma regimens with their monthly costs are shown in Table. 1 USD ≈ 154.39 ETB in early December 2025 [13].

The mean monthly cost of anti-glaucoma medication(s) was 293.40 ETB (range: 0–2610). The cost of IOP and visual field measurements was zero. The mean transportation cost was 444.37 ETB (range: 0–6000). The mean direct medical cost of all participants was 448.36 ETB (range: 0–5000), and the mean direct non-medical cost was 442.0 ETB (range: 0–6000). The mean total cost was 904.76 ETB (range: 30–6000). Direct costs constituted approximately 98% of the total cost (Table). 1 USD ≈ 154.39 ETB in early December 2025 [13].

Multiple linear regression analysis revealed that total cost was associated with residency, health insurance coverage, laterality of the disease, number of antiglaucoma medications, and history of glaucoma surgery. These associations were statistically significant with a p-value less than 0.05. Being from a rural area was associated with a 76.2% increase in total cost. Similarly, treatment of both eyes was associated with a 22.8% increase in the overall cost. Multidrug therapy was also associated with a higher cost burden than monotherapy by 208% and 64.8% for triple and dual therapy, respectively. Conversely, those with total and partial health insurance coverage of medication costs were associated with a 153.5% and 88.6% reduction in cost burden, respectively, compared with uninsured individuals. Having glaucoma surgery was also associated with a 29.6% decrease in total cost. However, the stage of glaucoma did not have a significant association with total cost burden (Table 4).

**Table 4:** The Monthly Costs of Anti-glaucoma Medications (in ETB)

| **Medications** | **Frequency (%)** | **Mean** | **SD** | **Min** | **Max** |
|---|---|---|---|---|---|
| Timolol | 232 (71.38%) | 82.54741 | 77.46401 | 0 | 500 |
| Timolol + Acetazolamide | 23 (7.08%) | 210.4348 | 175.3562 | 0 | 960 |
| Betaxolol | 1 (0.31%) | 300 | . | 300 | 300 |
| Timolol–Dorzolamide (fixed) + Acetazolamide | 2 (0.62%) | 655 | 77.78175 | 600 | 710 |
| Dorzolamide | 11 (3.38%) | 722.7273 | 271.444 | 0 | 950 |
| Timolol–Dorzolamide (fixed) | 7 (2.15%) | 727.1429 | 84.5999 | 600 | 850 |
| Timolol + Dorzolamide (fixed) | 29 (8.92%) | 796.5517 | 196.8008 | 0 | 1200 |
| Dorzolamide + Acetazolamide | 1 (0.31%) | 800 | . | 800 | 800 |
| Dorzolamide + Betaxolol | 1 (0.31%) | 1000 | . | 1000 | 1000 |
| Timolol + Pilocarpine | 5 (1.54%) | 1262 | 357.0994 | 860 | 1600 |
| Pilocarpine | 3 (0.92%) | 1500 | 100 | 1400 | 1600 |
| Timolol + Pilocarpine + Acetazolamide | 1 (0.31%) | 1860 | . | 1860 | 1860 |
| Pilocarpine + Timolol-Dorzolamide (fixed) | 2 (0.62%) | 1875 | 530.3301 | 1500 | 2250 |
| Timolol + Dorzolamide + Pilocarpine | 6 (1.85%) | 2130.833 | 264.3183 | 1640 | 2380 |
| Timolol + Dorzolamide + Latanoprost | 1 (0.31%) | 2610 | . | 2610 | 2610 |

**Table 4:**
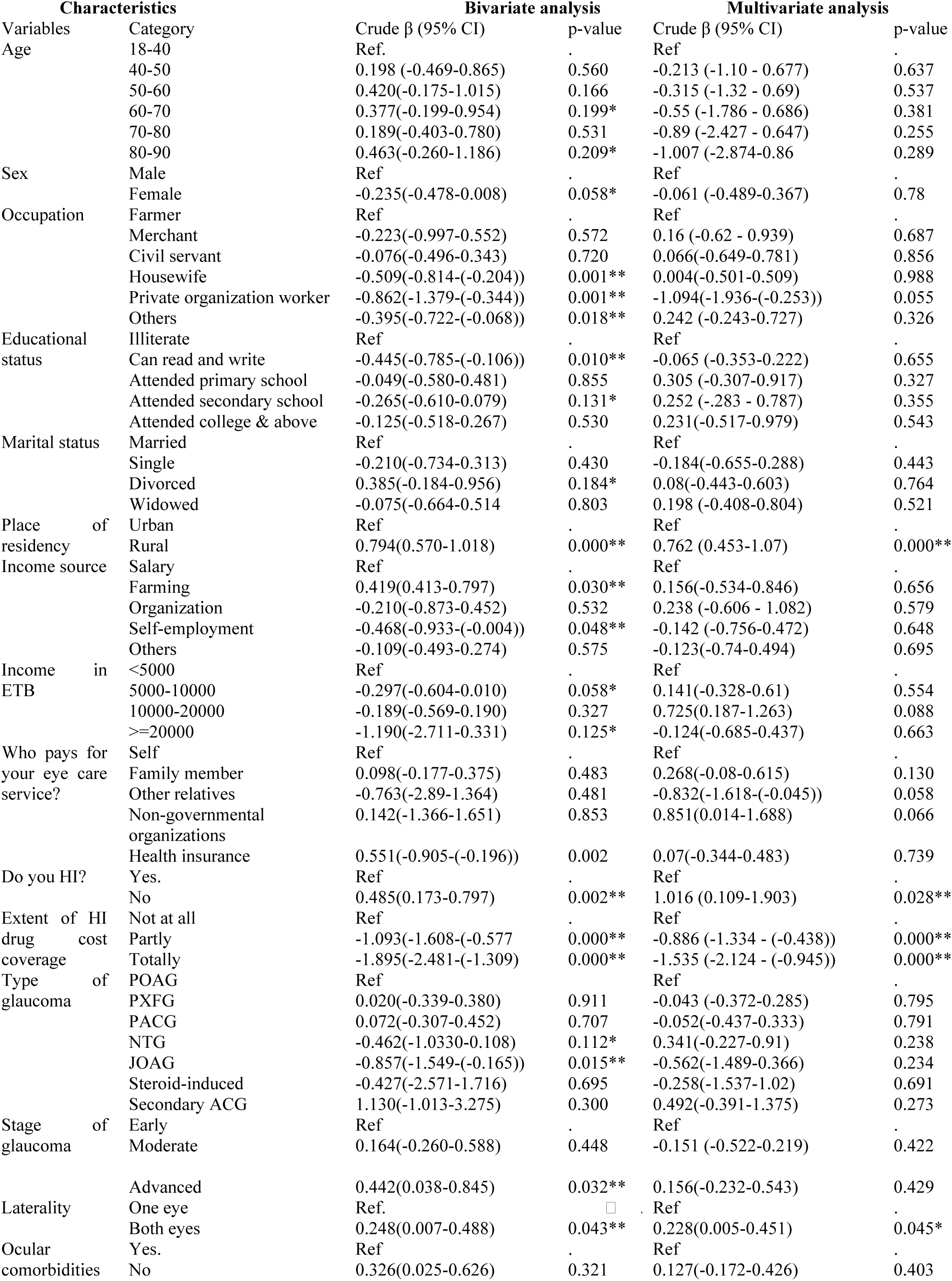
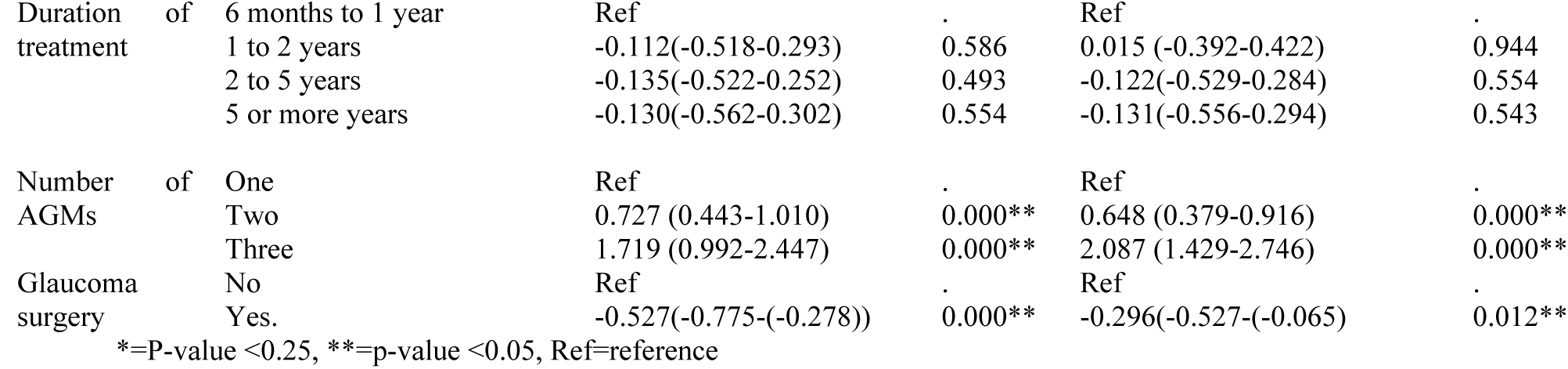
Linear Regression Analysis Results of Total Cost with Independent Variables.

| Characteristics |  | Bivariate analysis |  | Multivariate analysis |  |
|---|---|---|---|---|---|
| Variables | Category | Crude β (95% CI) | p-value | Crude β (95% CI) | p-value |
| Age | 18-40 | Ref. | . | Ref | . |
|  | 40-50 | 0.198 (-0.469-0.865) | 0.560 | -0.213 (-1.10 - 0.677) | 0.637 |
|  | 50-60 | 0.420(-0.175-1.015) | 0.166 | -0.315 (-1.32 - 0.69) | 0.537 |
|  | 60-70 | 0.377(-0.199-0.954) | 0.199* | -0.55 (-1.786 - 0.686) | 0.381 |
|  | 70-80 | 0.189(-0.403-0.780) | 0.531 | -0.89 (-2.427 - 0.647) | 0.255 |
|  | 80-90 | 0.463(-0.260-1.186) | 0.209* | -1.007 (-2.874-0.86 | 0.289 |
| Sex | Male | Ref | . | Ref | . |
|  | Female | -0.235(-0.478-0.008) | 0.058* | -0.061 (-0.489-0.367) | 0.78 |
| Occupation | Farmer | Ref | . | Ref | . |
|  | Merchant | -0.223(-0.997-0.552) | 0.572 | 0.16 (-0.62 - 0.939) | 0.687 |
|  | Civil servant | -0.076(-0.496-0.343) | 0.720 | 0.066(-0.649-0.781) | 0.856 |
|  | Housewife | -0.509(-0.814-(-0.204)) | 0.001** | 0.004(-0.501-0.509) | 0.988 |
|  | Private organization worker | -0.862(-1.379-(-0.344)) | 0.001** | -1.094(-1.936-(-0.253)) | 0.055 |
|  | Others | -0.395(-0.722-(-0.068)) | 0.018** | 0.242 (-0.243-0.727) | 0.326 |
| Educational status | Illiterate | Ref | . | Ref | . |
|  | Can read and write | -0.445(-0.785-(-0.106)) | 0.010** | -0.065 (-0.353-0.222) | 0.655 |
|  | Attended primary school | -0.049(-0.580-0.481) | 0.855 | 0.305 (-0.307-0.917) | 0.327 |
|  | Attended secondary school | -0.265(-0.610-0.079) | 0.131* | 0.252 (-.283 - 0.787) | 0.355 |
|  | Attended college & above | -0.125(-0.518-0.267) | 0.530 | 0.231(-0.517-0.979) | 0.543 |
| Marital status | Married | Ref | . | Ref | . |
|  | Single | -0.210(-0.734-0.313) | 0.430 | -0.184(-0.655-0.288) | 0.443 |
|  | Divorced | 0.385(-0.184-0.956) | 0.184* | 0.08(-0.443-0.603) | 0.764 |
|  | Widowed | -0.075(-0.664-0.514 | 0.803 | 0.198 (-0.408-0.804) | 0.521 |
| Place of residency | Urban | Ref | . | Ref | . |
|  | Rural | 0.794(0.570-1.018) | 0.000** | 0.762 (0.453-1.07) | 0.000** |
| Income source | Salary | Ref | . | Ref | . |
|  | Farming | 0.419(0.413-0.797) | 0.030** | 0.156(-0.534-0.846) | 0.656 |
|  | Organization | -0.210(-0.873-0.452) | 0.532 | 0.238 (-0.606 - 1.082) | 0.579 |
|  | Self-employment | -0.468(-0.933-(-0.004)) | 0.048** | -0.142 (-0.756-0.472) | 0.648 |
|  | Others | -0.109(-0.493-0.274) | 0.575 | -0.123(-0.74-0.494) | 0.695 |
| Income in ETB | <5000 | Ref | . | Ref | . |
|  | 5000-10000 | -0.297(-0.604-0.010) | 0.058* | 0.141(-0.328-0.61) | 0.554 |
|  | 10000-20000 | -0.189(-0.569-0.190) | 0.327 | 0.725(0.187-1.263) | 0.088 |
|  | >=20000 | -1.190(-2.711-0.331) | 0.125* | -0.124(-0.685-0.437) | 0.663 |
| Who pays for your eye care service? | Self | Ref | . | Ref | . |
|  | Family member | 0.098(-0.177-0.375) | 0.483 | 0.268(-0.08-0.615) | 0.130 |
|  | Other relatives | -0.763(-2.89-1.364) | 0.481 | -0.832(-1.618-(-0.045)) | 0.058 |
|  | Non-governmental organizations | 0.142(-1.366-1.651) | 0.853 | 0.851(0.014-1.688) | 0.066 |
|  | Health insurance | 0.551(-0.905-(-0.196)) | 0.002 | 0.07(-0.344-0.483) | 0.739 |
| Do you HI? | Yes. | Ref | . | Ref | . |
|  | No | 0.485(0.173-0.797) | 0.002** | 1.016 (0.109-1.903) | 0.028** |
| Extent of HI drug cost coverage | Not at all | Ref | . | Ref | . |
|  | Partly | -1.093(-1.608-(-0.577 | 0.000** | -0.886 (-1.334 - (-0.438)) | 0.000** |
|  | Totally | -1.895(-2.481-(-1.309) | 0.000** | -1.535 (-2.124 - (-0.945)) | 0.000** |
| Type of glaucoma | POAG | Ref |  | Ref | . |
|  | PXFG | 0.020(-0.339-0.380) | 0.911 | -0.043 (-0.372-0.285) | 0.795 |
|  | PACG | 0.072(-0.307-0.452) | 0.707 | -0.052(-0.437-0.333) | 0.791 |
|  | NTG | -0.462(-1.0330-0.108) | 0.112* | 0.341(-0.227-0.91) | 0.238 |
|  | JOAG | -0.857(-1.549-(-0.165)) | 0.015** | -0.562(-1.489-0.366) | 0.234 |
|  | Steroid-induced | -0.427(-2.571-1.716) | 0.695 | -0.258(-1.537-1.02) | 0.691 |
|  | Secondary ACG | 1.130(-1.013-3.275) | 0.300 | 0.492(-0.391-1.375) | 0.273 |
| Stage of glaucoma | Early | Ref | . | Ref | . |
|  | Moderate | 0.164(-0.260-0.588) | 0.448 | -0.151 (-0.522-0.219) | 0.422 |
|  | Advanced | 0.442(0.038-0.845) | 0.032** | 0.156(-0.232-0.543) | 0.429 |
| Laterality | One eye | Ref. | □ | Ref | . |
|  | Both eyes | 0.248(0.007-0.488) | 0.043** | 0.228(0.005-0.451) | 0.045* |
| Ocular comorbidities | Yes. | Ref | . | Ref | . |
|  | No | 0.326(0.025-0.626) | 0.321 | 0.127(-0.172-0.426) | 0.403 |
| Duration of treatment | 6 months to 1 year | Ref | . | Ref | . |
|  | 1 to 2 years | -0.112(-0.518-0.293) | 0.586 | 0.015 (-0.392-0.422) | 0.944 |
|  | 2 to 5 years | -0.135(-0.522-0.252) | 0.493 | -0.122(-0.529-0.284) | 0.554 |
|  | 5 or more years | -0.130(-0.562-0.302) | 0.554 | -0.131(-0.556-0.294) | 0.543 |
| Number of AGMs | One | Ref | . | Ref | . |
|  | Two | 0.727 (0.443-1.010) | 0.000** | 0.648 (0.379-0.916) | 0.000** |
|  | Three | 1.719 (0.992-2.447) | 0.000** | 2.087 (1.429-2.746) | 0.000** |
| Glaucoma surgery | No | Ref | . | Ref | . |
|  | Yes. | -0.527(-0.775-(-0.278)) | 0.000** | -0.296(-0.527-(-0.065) | 0.012** |

\*=P-value <0.25, \*\*=p-value <0.05, Ref=reference

### Intangible Burdens

As shown in Table 5, approximately 18.46% of patients quit or changed jobs due to glaucoma, and 10 (3.08) of the total participants stopped their education due to the disease. Seventy-eight (24%) patients experienced difficulties in performing routine activities that were easy to do previously. Of the total, 83 (25.54%) had difficulties in recognizing familiar faces, and 39 (12.00%) had difficulties in attending social events, such as weddings, ceremonies, and funerals.

**Table 5:**
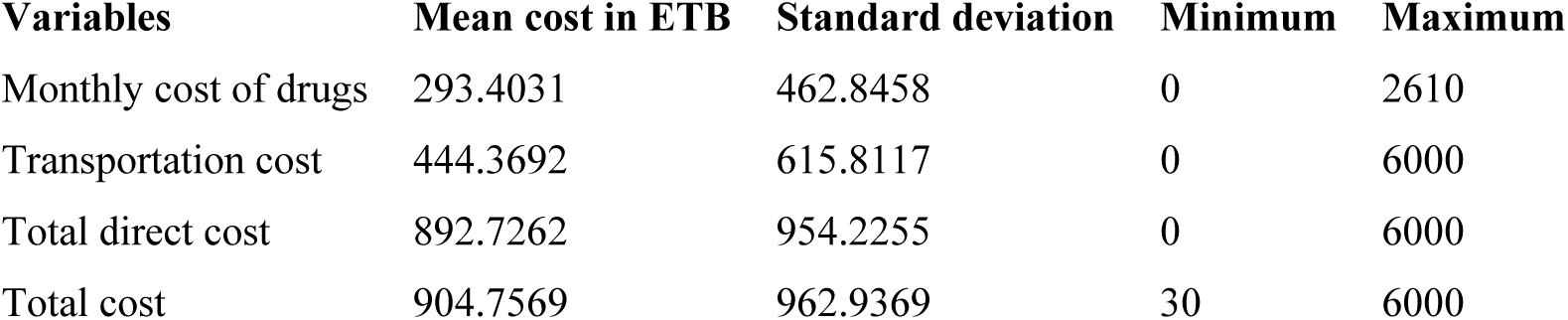
Intangible Burden-related Characteristics.

| **Variables** | **Mean cost in ETB** | **Standard deviation** | **Minimum** | **Maximum** |
|---|---|---|---|---|
| Monthly cost of drugs | 293.4031 | 462.8458 | 0 | 2610 |
| Transportation cost | 444.3692 | 615.8117 | 0 | 6000 |
| Total direct cost | 892.7262 | 954.2255 | 0 | 6000 |
| Total cost | 904.7569 | 962.9369 | 30 | 6000 |

**Table 5:** The Mean Costs, Including Total, Direct Costs, Medication, and Transportation Costs.

| **Characteristics** | **Category** | **Frequency (%)** |
|---|---|---|
| Patients quit or changed jobs. | Yes. | 60 (18.46%) |
|  | No | 265 (81.54%) |
| Patients quit education. | Yes. | 10 (3.08%) |
|  | No | 315 (96.92%) |
| Patients needed help from others because they were having difficulties performing routine activities. | Yes. | 78 (24%) |
|  | No | 247 (76%) |
| Patients had difficulties in recognizing familiar faces. | Yes. | 83 (25.54%) |
|  | No | 242 (74.46%) |
| Patients have difficulties in attending social events like weddings, funerals, ceremonies, etc. | Yes. | 39 (12%) |
|  | No | 286 (88%) |

### Discussions

Glaucoma is a critical and largely silent public health issue that disproportionately affects vulnerable, lower socioeconomic populations. The purchase of anti-glaucoma medication could pose a huge strain on patients’ income [4]. This study aimed to assess the socioeconomic burden of the disease and factors associated with higher financial distress among glaucoma patients visiting GUCSH, Northwest Ethiopia. The study identified that total glaucoma cost was significantly associated with residency, health insurance presence and extent of coverage, laterality of the disease, number of anti-glaucoma medications, and history of glaucoma surgery. These associations were statistically significant with a p-value less than 0.05.

The mean total cost related to glaucoma management in our study was 904.75 ETB (5.86 USD) per month, which was in the range of 0.2 to 38 USD. This constituted 22.1% of the mean monthly income. Direct costs constituted almost all (98%) of the total costs, and this was in accordance with a study conducted in Iran by Shakeri et al., which stated that direct costs constituted 94% of the total cost [14]. Moreover, the mean monthly cost of medications in this study was 293.40 ETB (range 0-2610 ETB), which was equivalent to 0 to 16 USD. Patients, on average, spent 7.2% of their monthly income on medication alone. This is comparable to a study done in Nigeria by Bello et al. (2023), which demonstrated that insured and noninsured patients spent 4.7% and 10.1% of their monthly income, respectively, on antiglaucoma medications [12]. A study in Jimma, Ethiopia, however, revealed that 29% of their income was spent on monthly antiglaucoma medications [4]. This discrepancy may be due to a difference in the proportion of participants who were on timolol alone, 71.38% in our study area and 40.5% in Jimma, respectively, as this drug was relatively cheaper and more easily available in public health facilities than other drugs. 1 USD ≈ 154.39 ETB in early December 2025 [13].

Based on our study, those who came from rural areas had a 76.2% increased total cost compared to urban dwellers, which was statistically significant with a p-value of 0.000. Similarly, a related study in Jimma, Ethiopia, also showed that patients who came from rural areas had a high direct cost compared with urban residents (1019.3 vs. 531.8 ETB), with a p-value of 0.001 [4]. The Ethiopian Birr and USD exchange rates were not mentioned. Our study also determined that treatment for both eyes had a 22% increased total cost (p=0.045) and a 29.7% increased medication cost (p=0.02) compared to treatment for one eye only. This finding is consistent with a study in the United States by Beal et al. (2021), which stated that payers of patients with bilateral disease had significantly higher annual costs for prescriptions. For at least one prescription in the bilateral cases, an additional $265 is needed annually (p<0.001) [15]. Similarly, the number of anti-glaucoma drugs was significantly associated with a higher cost burden than monotherapy by 208% for triple and 64.8% for dual therapy, with p-values of 0.000 and 0.000, respectively. The least costly medication in our study was timolol eye drops with a mean monthly cost of 82.5 ETB ($0.53), whereas the costliest one was triple therapy of timolol + dorzolamide + latanoprost with a monthly cost of 2610 ETB ($16.95). In line with this, a study in Nigeria reported that monotherapy with timolol was the least costly anti-glaucoma drug, with a mean monthly cost of Naira 640 ($1.8), whereas the three-drug combination of timolol, dorzolamide, and latanoprost was the costliest, at Naira 6761 ($18.8) [12].

Regarding health insurance influences, the study identified that having it with full and partial coverage of health care costs was significantly associated with 153.5% and 88.6% decrements in cost burden compared to uninsured individuals, with p-values of 0.01 and 0.031, respectively. Health insurance coverage in our study was 82.77%. Out of the insured groups, medicine expenses were covered fully (no additional out-of-pocket expenses for drugs) by the insurance in only 17.23%, partly in 78.81%, and not at all in 16 (5.95%) subjects by the time of the study. This parallels a study in Ghana in which 73.39% of the participants had health insurance coverage, and from these participants, 91% reported that the mere presence of it did not fully cover the expenses of their anti-glaucoma drugs [16]. Having glaucoma surgery was also significantly associated with a 29.6% decrement in total cost in our study (p-value = 0.012). Comparable to this, a study in Jimma, Ethiopia, reported that those who underwent surgery (trabeculectomy) had a non-statistically significant reduction in the cost of medication compared to those who did not (P=0.254) [4].

Although the total cost related to glaucoma in this study differed with the stage of glaucoma, the differences were not statistically significant. Conversely, studies in the United States and Argentina showed that increased severity of glaucoma was significantly associated with increased treatment costs, with p-values < 0.0001 and < 0.01, respectively [17, 18]. This discrepancy may be due to differences in disease-stage distribution and resulting statistical power; for instance, the majority of participants in our study were at the advanced stage (56.62%), as compared to these studies, in which mild glaucoma predominates (42.5% and 39.42%, respectively). This heavy concentration in the advanced group, combined with smaller sample sizes in the earlier stages, likely limited the statistical power needed to detect significant cost differences across groups.

Concerning intangible costs, our study showed that 18.46% of patients quit or changed jobs due to glaucoma; 24% of patients had difficulties in performing routine activities; 25.54% had difficulties in recognizing familiar faces; 12.00% had difficulties in attending social events such as weddings, ceremonies, and funerals; and 3.08% of participants stopped education. While those intangible burdens in a study conducted in Ghana align with our study, the magnitude of the effect differs. For instance, 50% of them stopped working, 83% had difficulties in performing routine tasks, and 19.33% had difficulties in attending social events [16]. This may be due to the difference in the stage of the disease at presentation or the presence of other visually significant comorbidities.

## Limitations

In this study, an accurate assessment of indirect costs was challenging, especially in estimating the cost of lost hours during follow-up visits. Accurate income assessment was also difficult, especially for those who described income in kind rather than in numbers. Furthermore, subjects may not genuinely report their actual income, which may affect our interpretation.

## Conclusion and Recommendations

Glaucoma is a progressive disease; hence, it necessitates lifelong treatment. This study identified that the management of glaucoma had a significant effect on patients’ economic and/or personal as well as social quality of life. Our patients spent 22.1% of their reported monthly income on the total costs of glaucoma. Patients from rural areas spent more. Those who were on treatment for both eyes were also victims of a higher financial burden. Health insurance, despite its limitations on full functionality, significantly reduced total and direct cost burdens. Having glaucoma surgery has also been shown to reduce the total cost, as it alters the number of medications and follow-up durations. Glaucoma-related visual impairment can lead to personal and social incapacitation; this, in turn, may pose a financial burden. We recommend that broader health insurance coverage for more anti-glaucoma medications, including those bought from private pharmacies, and an uninterrupted supply of common anti-glaucoma medications in public health settings could decrease the economic burden to patients. It is better to consider surgical options earlier, especially for those who come from distant areas, as it can reduce financial constraints. The government, health system, and stakeholders should stand together to educate the community as a whole, especially older at-risk groups, to have earlier glaucoma screening, start timely management, address disabilities from glaucoma, and reduce economic handicaps.

## List of Abbreviations and Acronyms

ACG: angle closure glaucoma
AGM: anti-glaucoma medications
AMD: age-related macular degeneration
ANOVA: analysis of variance
ARIC: age-related immature cataract
C/D*: Cup to disk ratio
COI: cost of illness
DM: diabetes mellitus
ETB: Ethiopian Birr
GUCSH: Gondar University Comprehensive Specialized Hospital
HI: health insurance
HTN: hypertension
IOP: intraocular pressure
MD: mean deviation
MGD: meibomian gland dysfunction
NGO: Nongovernmental Organization
NTG: normal tension glaucoma
OAG: open-angle glaucoma
ONH: optic nerve head
PACG: primary angle closure glaucoma
PAC: primary angle closure
POAG: primary open-angle glaucoma
PUD: peptic ulcer disease
RVO: retinal vein occlusion
UOG: University of Gondar
USD: United States Dollar
VIF: Variance Inflation Factor
VKC: Vernal keratoconjunctivitis
WHO: World Health Organization

## Data Availability

The de-identified dataset underlying this study will be deposited in the Zenodo repository and made publicly available. The DOI will be provided before publication, subject to confirmation that data sharing complies with the study's ethical approval and applicable institutional requirements.

## Acknowledgment

We would like to thank the University of Gondar, College of Medicine and Health Science, Department of Ophthalmology, and the Tertiary Eye Care and Training Center for allowing us to conduct this study. We are also truly grateful to the study participants and colleagues who played a tremendous role during the data collection process.

## Authors’ contributions

**Conceptualization**: Fisseha Admassu.

**Data curation**: Simegnew Nibret

**Methodology**: Banchilay Addis, Tesfahun Ejigu.

**Investigation**: Getasew Ayichew, Simegnew Nibret.

**Formal analysis**: Banchilay Addis, Simegnew Nibret.

**Writing** – **original draft**: Simegnew Nibret.

**Writing – review & editing**: Fisseha Admassu, Tesfahun Ejigu, Banchilay Addis.

**Supervision**: Fisseha Admassu, Tesfahun Ejigu, Banchilay Addis.

## Declarations

### Ethical approval and consent to participate

Ethical approval was obtained from the University of Gondar School of Medicine Ethical Review Committee (Reference No. SOM 460/2025). Written informed consent was obtained from all participants before enrollment in the study.

